# Leveraging Large Language Models for Digital Phenotyping: Detecting Depressive State Changes for Patients with Depressive Episodes

**DOI:** 10.1101/2025.05.10.25327354

**Authors:** Yunhao Yuan, Ya Gao, Hans Moen, Erkki Isometsä, Pekka Marttinen, Talayeh Aledavood

## Abstract

Digital phenotyping, which takes advantage of data continuously gathered from smartphones and wearable devices, offers promising avenues for real-time monitoring and mental health analysis. This approach holds promise for improving early detection and personalized care in mood disorders by enabling clinicians to proactively respond to significant changes before symptoms worsen. However, the complexity and heterogeneity of digital phenotyping data pose significant modeling challenges. Recent advances in large language models (LLMs) suggest their potential to generalize across diverse tasks with minimal labeled data, making them a promising alternative for analyzing data from digital phenotyping studies. However, the extent of usability of these methods for digital phenotyping studies is not yet well understood. In this study, we evaluate the potential of LLMs in analyzing digital phenotyping data to predict changes in depression severity among individuals experiencing major depressive episodes. We evaluate several in-context learning and fine-tuning strategies, and find that both few-shot prompted LLMs and fine-tuned models outperform traditional machine learning baselines trained on the same set of input features. Moreover, we compare two fine-tuning approaches (fine-tuning only the embedding layer versus parameter-efficient fine-tuning using QLoRA) and find that fine-tuning only the embedding layer significantly improves performance compared to QLoRA fine-tuning. These results highlight the capability of LLMs to process and integrate heterogeneous behavioral data, promising their application for digital phenotyping and mental health research. While our findings highlight the potential in using LLMs in mental health monitoring, their black-box nature and risk of replicating data biases highlight the need for clinical oversight and validation in real-world practice.

## 1 Introduction

Recent years have seen increasingly rapid advances in large language models (LLMs), including notable examples such as ChatGPT-4 [1], LLaMA-3.1 [2], and DeepSeek [3]. These models are capable of encoding vast amounts of information and demonstrate robust performance across diverse tasks involving complex natural language understanding, reasoning, and generation, such as question answering, summarization, and problem solving [4–7]. In the health domain, there is also a growing interest in utilizing LLMs to enhance clinical decision-making, electronic health records analysis, patient communication, medical education, and medical images interpretation and assessments [8–14]. Despite these promising developments, relatively few studies have explored the potential of LLMs within the emerging field of digital phenotyping.

Digital phenotyping refers to the quantification of human behaviors, physiological states, and clinical characteristics using data collected passively and continuously from smartphones, wearables, and other connected devices [15–19]. By constructing detailed digital trajectories of patients’ behavioral, social, and physiological patterns, this approach offers a dynamic and personalized window into mental and physical health [20]. Digital phenotyping studies often extract behavioral markers from passively collected data and find associations between them and self-reported questionnaires, which, for example, assess symptoms of depression. The behavioral markers are also used to predict future questionnaire scores or changes in them. Past studies have largely relied on linear regression models [21, 22] and non-linear machine learning models [23–26] for these goals. However, these methods often struggle with the scalability and adaptability required to manage the complexity of real-world digital data [27].

Recent advances in deep learning have opened up new possibilities for addressing these challenges, yet their implementation in digital phenotyping remains limited by the need for substantial amounts of clinician-annotated data to achieve robust performance [28]. Interestingly, LLMs demonstrate exceptional generalizability. Pre-trained on hundreds of billions of tokens from diverse public corpora, LLMs can perform unseen tasks with little or no task-specific data through zero- and few-shot in-context learning, where behavior is guided by a few examples or even a single prompt [29].

When limited labeled data is available, the fine-tuning techniques [30] can further specialize LLMs for clinical applications with minimal annotation effort. Together, incontext learning and fine-tuning enable LLMs to achieve strong performance in digital phenotyping tasks, even under conditions of limited supervision. In this work, we evaluate an open-source LLM (Llama 3.1-Instruct-8B) on digital phenotyping data. Specifically, we address the following research question: Can large language models accurately detect changes in depression severity using digital phenotyping data?

We use a digital phenotyping dataset [20, 31, 32] collected via smartphone applications from patients with major depressive episodes and healthy controls recruited from multiple clinics across Finland. Our evaluation framework evaluates the ability of LLM to predict the changes in self-reported depression severity through digital phenotyping. The initial depression severity of a participant is determined based on the Patient Health Questionnaire (PHQ-9), which asks about their state over the past two weeks. The task of the LLM is then to detect whether a participant’s depressive state changes from one two-week period to the subsequent two-week period using passively collected digital phenotyping data. We conduct a series of experiments using Llama 3.1-Instruct-8B to detect the changes in depression severity based on passively collected digital phenotyping data. We explore and compare various prompting strategies and fine-tuning techniques to analyze the LLM’s capabilities in digital phenotyping research. We also compare this to a more traditional classification model, XGBoost, which has demonstrated strong performance in prior digital phenotyping studies [20].

## 2 Results

### 2.1 Study cohort and study design

We analyze data from the Mobile Monitoring of Mood (MoMo-Mood) study [20], a 1-year digital phenotyping study of 133 participants who have a diagnosis of either major depressive disorder (MDD) (mean age: 39.0, SD: 14.2), major depressive episodes (MDE) with bipolar disorder (BD) (mean age: 37.1, SD: 10.3), or MDD with borderline personality disorder (BPD) (mean age: 28.3, SD: 6.0) and 31 healthy controls (mean age: 41.8, SD: 13.9).

The specific prediction task requires the LLMs to classify future depressive state changes (increase, decrease, or stable) based on smartphone-derived digital phenotyping data, initial depressive states, and optional historical context. The structure of the prompt given to the LLM is shown in Figure 2. Participants’ mobile sensor data—including battery level, screen usage, acceleration magnitude, and screen-off durations—are collected continuously and paired with biweekly PHQ-9 assessments [33], a 9-item clinical questionnaire that evaluates the severity of depressive symptoms, with higher scores indicating more severe depression (Figure 1). These passively collected behavioral signals can be used as proxies for mental health indicators such as social withdrawal, disrupted sleep, or reduced mobility, which are associated with depressive episodes [20]. We reformulate the detection of depressive state changes as a natural language task, enabling the application of LLMs to this digital phenotyping dataset.

**Fig. 1.**
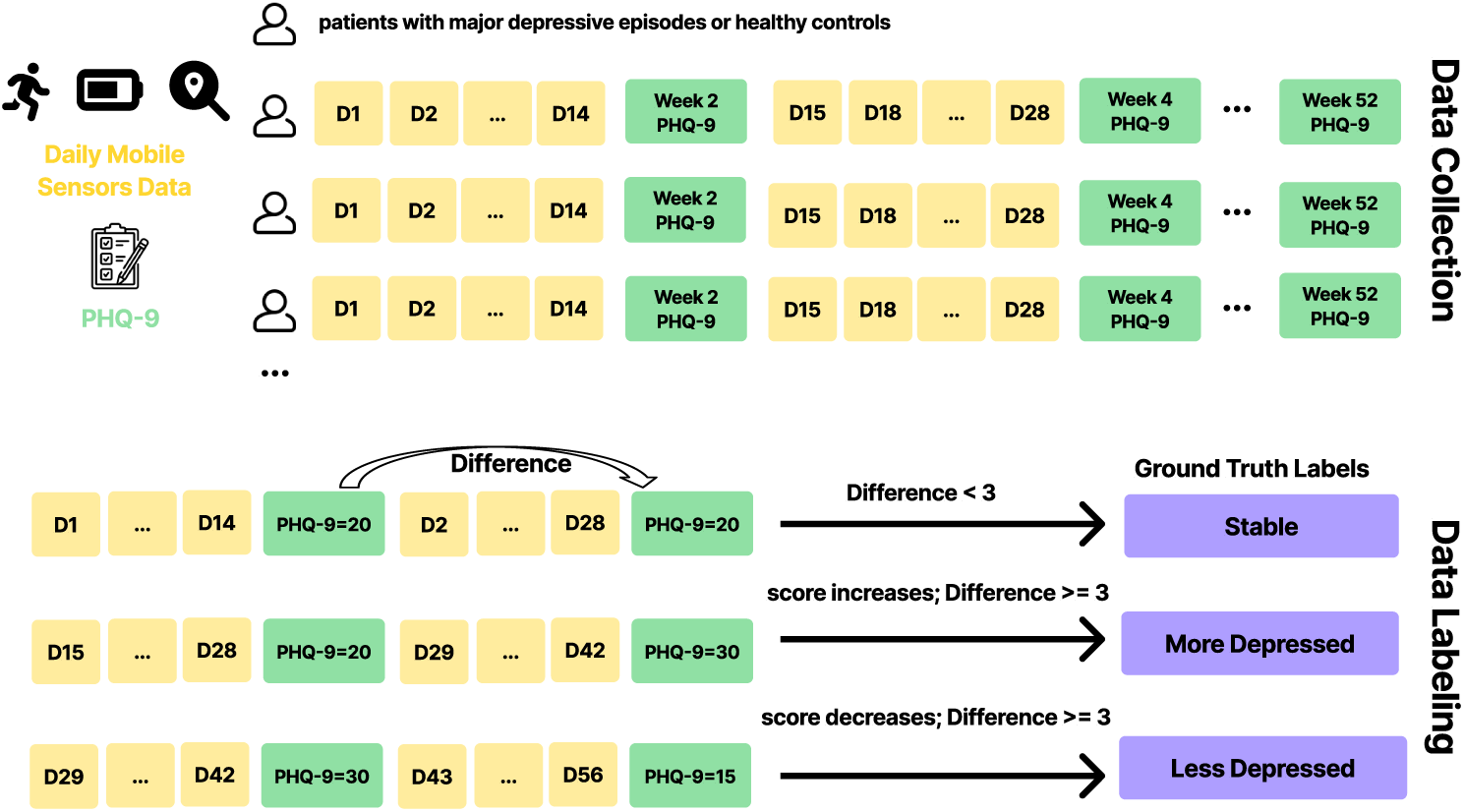
Data Collection and Labeling Process. The upper panel illustrates data collection from subjects during the passive phase, where daily phenotypic data (D1, D2, etc.) and bi-weekly depression levels (PHQ-9 scores) are recorded. The lower panel depicts the data labeling process: ground truth labels are assigned based on the difference in PHQ-9 scores between two consecutive two-week periods. These labels indicate whether an individual’s depression severity remains stable, increases, or decreases across the compared periods.

**Fig. 2.**
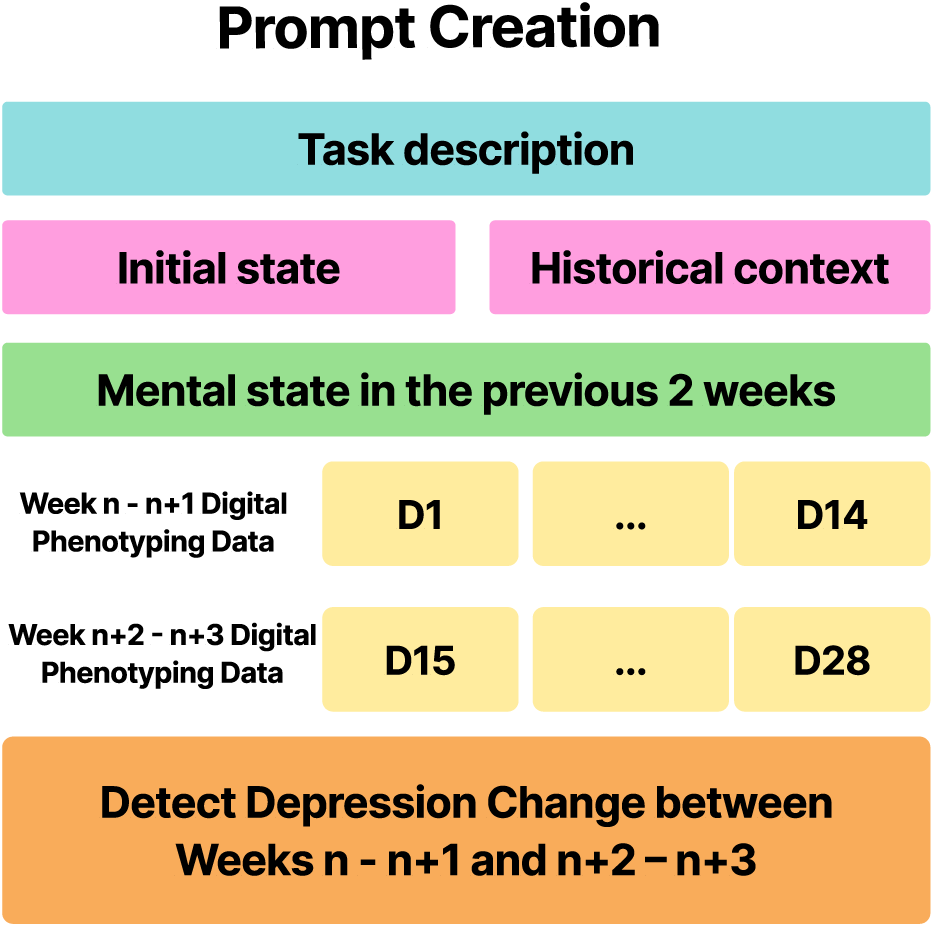
Our structured prompt consists of a task description, participants’ initial mental state, optional historical context, daily behavioral and mental state data spanning two consecutive 2-week periods (D1–D14 and D15–D28, where D indicates the day index within two consecutive 2-week period), and an instruction sentence prompting the model to predict changes in depressive state between these two periods. The predicted label corresponds to the change in mental state over time (e.g., increase or decrease in depressive symptoms). An example of the prompt is shown in Table 2.

We use two modeling strategies: few-shot in-context learning and supervised finetuning. To evaluate model performance under diverse conditions, we explore 288 unique configurations resulting from combinations of modeling strategy, input features, prompt format, and training setup. Specifically, we test 16 digital phenotyping feature combinations (battery usage, screen-off duration, screen-use duration, and acceleration magnitude) under two prompt formats: *init* (includes the participant’s initial depressive state and digital phenotyping data from the most recent four-week period;) and *init history* (extends the *init* format by adding a summary of the participant’s behavioral context and mental state from the prior two-week period). For in-context learning, we evaluate seven few-shot settings (0, 1, 3, 5, 10, 15, and 25 examples), resulting in 224 configurations. For fine-tuning, we compare two strategies: parameter-efficient fine-tuning using QLoRA [34] and embedding-layer-only fine-tuning, across the same 16 feature sets and two prompt formats, yielding 64 additional configurations. All experiments are repeated across five random seeds to ensure the robustness of the results. We evaluate model performance using average Macro F1 scores (Tables 1.

We compare the LLM-based models against an XGBoost baseline trained on the same input features as LLMs. XGBoost is selected as a comparison method because it has demonstrated strong performance in prior digital phenotyping studies focused on predicting depression severity and mental health outcomes using a digital phenotyping dataset [20].

### 2.2 In-Context Learning Performance

Table 1 represents the results of our in-context learning experiments. In this approach, the model relies entirely on the provided task-specific context, numerical values, and a limited number of few-shot examples to detect depression changes without updating the parameters within the pre-trained model. All performance metrics are averaged over five independent runs with different random seeds, ensuring the robustness of our findings. By evaluating in-context learning, we assess the extent to which large language models can generalize to digital phenotyping tasks without the need for computationally expensive retraining on labeled data.

**Table 1.** Performance comparison of in-context learning and fine-tuning strategies with different feature sets for detecting depressive state changes. The table presents Macro F1 scores (averaged over five independent runs with different random seeds) for different combinations of digital phenotyping features, and a comparison with the XGBoost baseline. Two prompt formats are tested: *init*, which includes only initial depressive state indicators and four weeks of digital phenotyping behavioral data, and *init history*, which incorporates initial depressive state, historical behavioral context, and four weeks of digital phenotyping behavioral data.

| Feature | Prompt | Few-shots learning |  |  |  |  |  |  | Fine-tuning |  | XGboost |
| --- | --- | --- | --- | --- | --- | --- | --- | --- | --- | --- | --- |
|  |  | 0 | 1 | 3 | 5 | 10 | 15 | 25 | QLora | Embedding |  |
| Battery | init | 0.220 | 0.288 | 0.281 | 0.298 | 0.311 | 0.322 | **0.343** | 0.294 | 0.327 | 0.307 |
|  | init.history | 0.250 | 0.307 | 0.264 | 0.297 | 0.307 | 0.326 | **0.344** | 0.294 | 0.319 |  |
| Screen-Off | init | 0.330 | 0.318 | 0.314 | 0.314 | **0.345** | 0.330 | 0.340 | 0.288 | 0.338 | 0.352 |
|  | init.history | 0.355 | 0.293 | 0.320 | 0.319 | 0.341 | 0.352 | **0.361** | 0.296 | 0.345 |  |
| Screen-Use | init | 0.245 | 0.289 | 0.264 | 0.269 | 0.325 | 0.317 | **0.346** | 0.300 | 0.315 | 0.325 |
|  | init.history | 0.240 | 0.268 | 0.275 | 0.291 | 0.340 | 0.336 | **0.355** | 0.292 | 0.330 |  |
| Magnitude | init | 0.281 | 0.314 | 0.290 | 0.279 | 0.305 | 0.317 | 0.312 | 0.320 | 0.334 | 0.324 |
|  | init.history | 0.339 | 0.323 | 0.303 | 0.327 | 0.342 | **0.348** | 0.334 | 0.294 | 0.321 |  |
| Battery<br>+ Screen-Off | init | 0.237 | 0.314 | 0.261 | 0.297 | 0.323 | 0.320 | **0.345** | 0.294 | 0.301 | 0.303 |
|  | init.history | 0.206 | 0.313 | 0.262 | 0.290 | 0.338 | 0.326 | **0.351** | 0.294 | 0.306 |  |
| Battery<br>+ Screen-Use | init | 0.242 | 0.315 | 0.256 | 0.282 | 0.312 | 0.310 | **0.336** | 0.294 | 0.336 | 0.294 |
|  | init.history | 0.246 | 0.317 | 0.268 | 0.282 | 0.335 | 0.331 | **0.350** | 0.294 | 0.329 |  |
| Screen-Off<br>+ Screen-Use | init | 0.276 | 0.321 | 0.286 | 0.301 | 0.359 | **0.364** | 0.357 | 0.307 | 0.311 | 0.313 |
|  | init.history | 0.258 | 0.318 | 0.292 | 0.309 | 0.362 | 0.372 | **0.378** | 0.301 | 0.327 |  |
| Battery<br>+ Magnitude | init | 0.273 | 0.303 | 0.252 | 0.278 | 0.322 | 0.330 | **0.340** | 0.294 | 0.306 | 0.292 |
|  | init.history | 0.234 | 0.319 | 0.269 | 0.303 | 0.355 | 0.339 | **0.359** | 0.291 | 0.319 |  |
| Screen-Off<br>+ Magnitude | init | 0.260 | 0.347 | 0.307 | 0.320 | 0.340 | **0.360** | 0.356 | 0.324 | 0.294 | 0.332 |
|  | init.history | 0.295 | 0.352 | 0.309 | 0.336 | 0.348 | 0.340 | **0.375** | 0.323 | 0.327 |  |
| Screen-Use<br>+ Magnitude | init | 0.223 | 0.309 | 0.290 | 0.305 | 0.340 | 0.329 | **0.353** | 0.294 | 0.318 | 0.329 |
|  | init.history | 0.216 | 0.313 | 0.291 | 0.326 | 0.335 | 0.352 | **0.351** | 0.294 | 0.331 |  |
| Battery<br>+ Screen-Off<br>+ Screen-Use | init | 0.225 | 0.321 | 0.303 | 0.332 | 0.338 | 0.324 | **0.356** | 0.294 | 0.309 | 0.297 |
|  | init.history | 0.208 | 0.302 | 0.259 | 0.295 | 0.346 | 0.331 | **0.356** | 0.291 | 0.296 |  |
| Magnitude<br>+ Screen-Off<br>+ Screen-Use | init | 0.231 | 0.303 | 0.263 | 0.29 | 0.339 | 0.331 | **0.402** | 0.298 | 0.311 | 0.305 |
|  | init.history | 0.297 | 0.302 | 0.249 | 0.275 | 0.346 | 0.336 | **0.396** | 0.321 | 0.305 |  |
| Battery<br>+ Magnitude<br>+ Screen-Use | init | 0.228 | 0.309 | 0.283 | 0.322 | 0.335 | 0.340 | **0.359** | 0.292 | 0.308 | 0.295 |
|  | init.history | 0.245 | 0.307 | 0.241 | 0.313 | 0.339 | 0.342 | **0.371** | 0.292 | 0.315 |  |
| Battery<br>+ Screen-Off<br>+ Magnitude | init | 0.216 | 0.301 | 0.296 | 0.316 | 0.344 | 0.336 | **0.362** | 0.293 | 0.313 | 0.297 |
|  | init.history | 0.227 | 0.299 | 0.256 | 0.309 | 0.345 | 0.351 | **0.355** | 0.294 | 0.306 |  |
| All Features | init | 0.289 | 0.311 | 0.260 | 0.290 | 0.332 | 0.336 | **0.377** | 0.292 | 0.305 | 0.308 |
|  | init.history | 0.298 | 0.331 | 0.275 | 0.330 | 0.360 | 0.361 | **0.364** | 0.292 | 0.303 |  |

**Table 2.** Example of a structured prompt used for in-context learning and fine-tuning in detecting depressive state changes.

| Prompt |  | Input |
| --- | --- | --- |
| **Role** |  | You are an intelligent healthcare agent skilled in detecting changes in mental health conditions. |
| **Task** |  | Your task is to determine how the user’s mental health state changes based on digital data of mobile sensor readings from an individual. The input contains the following contents: data about smartphone usage over the previous two weeks, whether being depressed or not over the previous two weeks, and data about smartphone usage over the current two weeks. To determine how the user’s mental health state changes, you should only respond with "Label: Remains stable" or "Label: More Depressed" or "Label: Less Depressed". |
| **Con-**<br>**text** | **Initial state** | The following data come from a {50-year-old} female. This person was *not depressed* when they first entered the experiment. NaN denotes missing data. |
|  | **Historical Context** | Within the 2 weeks so far, the average smartphone usage: *acceleration magnitude*: {9.9}. |
|  | **Previous 2 weeks Data and Mental state** | This person was in {*not depressed*} over the previous two-week. Smartphone over the previous two weeks: Max values of the acceleration magnitude within the day over the previous two weeks, measured using the accelerometer of the phone: {9.9, ..., 9.3} |
|  | **Current 2 Weeks Data** | smartphone data over the current two-week: Max values of the acceleration magnitude within the day over the current two-week, measured using the accelerometer of the phone: {9.3, ..., 9.3} |
| **Output** |  | Determine how the mental health state changes and only output the label: |

Our findings reveal that the pre-trained LLM can effectively identify meaningful patterns in the digital phenotyping data, even with limited examples. Notably, utilizing only screen usage duration in the afternoon sequence and initial depressive state indicators *init*, the average Macro F1 score improves from 0.245 in the zero-shot setting to 0.346 in the 25-shot setting, highlighting the model’s ability to generalize from limited examples. Similarly, when the feature set is expanded to *Magnitude + Screen-Use + Screen-off* with initial depressive state indicators *init*, the model achieved its highest performance of 0.402 in the 25-shot setting. This highlights the LLM’s ability to integrate multiple features, enhancing the performance of detecting depressive state transitions.

Our results also show that few-shot in-context learning outperforms both zero-shot and the XGBoost baseline [35]. For instance, under the *init history* prompt, 25-shot in-context learning achieves a Macro F1 score of 0.377 with the “All Features” set—substantially higher than the 0.308 achieved by XGBoost trained on labeled training data with the same features. Similarly, selecting targeted features including *Battery, Screen-off,* and *Magnitude* yields even larger improvements, where 25-shot prompting surpasses the XGBoost baseline by over 0.058 points in F1. This demonstrates that LLMs can learn contextual and sequential information in the dataset, often outperforming traditional machine learning models. Also, this performance advantage is achieved with far less labeled training data. XGBoost requires access to the full training dataset to learn its parameters—amounting to 50% of our dataset. In contrast, in-context learning models perform competitively with only a small number of examples (e.g., 25-shot prompting uses just 25 labeled examples), showcasing the data efficiency of LLMs when supplied with well-structured task prompts.

Interestingly, while the 1-shot example generally outperforms the zero-shot scenario, the 3-shot and 5-shot examples often show slightly lower performance than the 1-shot setting. For example, when using screen-off duration during the night and maximum acceleration magnitude (*Screen-off + Magnitude*) with an initial depressive indicator and historical behavioral context (*init history*) as the input feature, the F1 score decreases from 0.352 (1-shot) to 0.309 (3-shot) and 0.336 (5-shot) before rising again to 0.375 in the 25-shot setting. This trend, observed across multiple feature combinations, suggests that the model may initially struggle to adapt and generalize as additional, limited examples are introduced, but benefits significantly from larger sets of training examples (e.g., 10-shot and 25-shot scenarios).

### 2.3 Fine-Tuning Performance

Table 1 presents the results of our fine-tuning experiments for detecting depressive state changes using digital phenotyping data. In this approach, the model undergoes parameter updates to adapt its learned representations to the specific task, allowing it to internalize and refine patterns from digital phenotyping data. Two distinct fine-tuning strategies—parameter-efficient fine-tuning using QLoRA and embedding layer fine-tuning are compared across diverse feature sets to determine their effectiveness in capturing behavioral patterns associated with depressive state transitions. By analyzing fine-tuning performance, we aim to assess how well LLMs can learn from digital phenotyping data when given access to labeled training samples and optimized through targeted parameter adjustments.

We also find that fine-tuning the embedding layer outperforms QLoRA fine-tuning in capturing complex human behavioral patterns, particularly when combined with historical behavioral context (*init history*). Fine-tuning the embedding layer shows average Macro F1 improvements in classification performance across most feature sets. For example, using the sequences of screen-off duration at night, embedding layer fine-tuning achieves an average Macro F1 score of 0.345 with the initial depressive status and historical summaries in the prompt (*init history*), compared to 0.294 with QLoRA fine-tuning. Similarly, in the average battery level in the afternoon feature set with initial depressive status (*init*), the Macro F1 score increases from 0.294 (QLoRA fine-tuning) to 0.327 (embedding-layer fine-tuning). We also notice that QLoRA finetuning achieves a Marco F1 score of around 0.29 across most configurations. This lack of variation suggests that QLoRA fine-tuning may lead to overfitting.

In addition, examining the impact of incorporating historical depressive status information (*init history*) versus using only the initial status (*init*) highlights further nuances. When restricting the model to screen-off durations, the Macro F1 improves from 0.338 (*init*) to 0.345 (*init history*) in the embedding layer fine-tuning, emphasizing that knowledge of a user’s prior depressive state can enhance classification accuracy. In contrast, the benefit of historical context is less pronounced—or even slightly detrimental—for other feature sets, such as *Magnitude Only* (0.334 vs. 0.321) and *Battery + Magnitude + Screen-Use* (0.308 vs. 0.315). These disparities suggest that the utility of historical context may depend on how strongly a particular feature (e.g., night screen-off patterns) correlates with shifts in depressive states, thus making historical data more directly relevant for certain behaviors compared to others. The generally lower scores under QLoRA fine-tuning—even when multiple features are combined—reinforce the observation that adjusting only the embedding layer can help avoid overfitting while still allowing meaningful refinement of behavioral cues for depressive state detection.

When comparing fine-tuning to the XGBoost baseline, we observe that embedding-layer fine-tuning often performs comparably or better across several feature sets, especially when using richer behavioral information. For instance, when using *Battery* feature with *init* prompts, embedding-layer fine-tuning achieves an average Macro F1 of 0.327, which is slightly better than the XGBoost score of 0.307. However, in combinations that include more contextual behavioral signals—such as *Battery + Screen-Use* with *init* prompts (0.336 vs. 0.294) or *Battery + Magnitude + Screen-Use* with *init history* prompts (0.315 vs. 0.295)—embedding fine-tuning frequently exceeds XGBoost performance. Both fine-tuning and XGBoost are trained on the same full training dataset; however, when compared to few-shot in-context learning, embedding-layer fine-tuning often performs lower, even though in-context learning uses significantly less labeled data (only 25 examples embedded in the prompt). This suggests that few-shot in-context learning is not only highly data-efficient but also highly competitive, demonstrating that large language models can generalize well from limited, well-structured examples without the need for parameter updates.

## 3 Discussion

We develop LLM-based multi-label classifiers from digital phenotyping data to detect changes in depressive state. By leveraging in-context learning and fine-tuning strategies, we show that LLMs can integrate complex behavioral features to detect mental health outcomes better than the machine learning baseline–trained on labeled training data with the same feature sets. These results suggest the promising future of using LLM in mental health monitoring tasks.

### 3.1 In-Context Learning For Digital Phenotyping Data

Our experiments with in-context learning reveal that LLMs can effectively analyze digital phenotyping data using their pre-trained knowledge. In-context learning allows models to make detections without additional training by embedding task-specific examples directly into the input prompt [36]. This method utilizes the model’s vast pre-existing knowledge, which shows a significant advantage for real-time deployment in mental health monitoring applications.

However, the performance of in-context learning varies depending on the number and quality of examples provided. In zero-shot scenarios, where the LLM relies entirely on its pre-existing capabilities, performance tends to be lower. When a single example is introduced, performance improves significantly, showing that even minimal guidance can help the model understand the task context. On the other hand, adding additional but still limited examples (e.g., 3- or 5-shot settings) sometimes results in decreased performance. This transient decline suggests that noisy or inconsistent examples may confuse the model, underscoring the importance of prompt engineering.

When the number of examples increases beyond 5-shot, e.g., 10 or 25 examples, performance improves consistently, indicating that a larger number of examples provides greater clarity and enables the model to generalize better. These results seem to indicate that there is a trade-off between having the LLM primarily rely on inbuilt prior knowledge versus trying to generalize from in-context examples. In the latter case, the coverage, amount, and quality of the examples become especially important. Further, these results highlight the inherent strengths of LLMs in pattern extraction and their reliance on high-quality, structured input examples, especially in tasks involving varied behavioral contexts. We see the task of further optimizing what in-context examples and information to present to the LLM as a natural direction for future work. This includes exploring retrieval-augmented generation (RAG) approaches [37, 38], where the objective would be to provide contextually dependent input examples optimized for each classification instance.

Unlike more traditional machine learning models, which typically require explicit feature engineering to capture temporal dependencies, LLMs can recognize patterns directly from sequences of digital phenotyping data. This suggests that, with the appropriate prompts, LLMs can act as powerful tools for analyzing mental health-related behaviors. However, as seen in our results, the variability in performance across few-shot settings reinforces the need for continued exploration into the best strategies for prompt optimization, particularly in mental health-related tasks, where participant behaviors and contexts are often complex and diverse.

### 3.2 Fine-Tuning Strategies

In addition to in-context learning, we assess two different fine-tuning strategies (parameter-efficient fine-tuning using QLoRA and embedding layer fine-tuning). Our findings reveal that embedding-layer fine-tuning generally outperforms QLoRA fine-tuning across multiple feature sets and prompt formulations.

Embedding-layer fine-tuning proves to be a more effective strategy, primarily because it updates only the input embeddings during training. This approach preserves the foundation knowledge embedded in the deeper layers of the pre-trained model while adapting its representation space to task-specific data. In contrast, QLoRA model fine-tuning, which adjusts all parameters, can lead to overfitting and a loss of generalizability. This issue is particularly pronounced in mental health applications, where the data is often sparse, noisy, and subject to significant individual variability, making it crucial to retain the foundational knowledge of the pre-trained model.

The computational efficiency of embedding-layer fine-tuning is another key advantage. Reducing the number of parameters that need to be updated reduces the computational overhead, offering a more practical solution for resource-constrained environments. This is particularly relevant in the context of mental health monitoring, where real-time, low-cost, and scalable solutions are essential.

### 3.3 Comparing In-Context Learning and Fine-Tuning

While both in-context learning and fine-tuning show strong classification capabilities, their performance differs [39]. In-context learning, particularly with a larger set of fewshot examples, consistently demonstrates strong performance, effectively integrating multiple behavioral features without relying heavily on historical behavioral context. In contrast, embedding-layer fine-tuning revealed nuanced sensitivity to historical depressive information in the context. Specifically, incorporating historical context enhances performance notably for certain features, such as screen-off durations, indicating the fine-tuned model’s capability to use prior depressive state information to better interpret current behavior [40, 41]. However, this advantage is less consistent across other combinations of behavioral features, such as when using *magnitude* feature as input, suggesting that the value of historical context is conditional upon the strength of the relationship between the behavioral and depressive states within specific feature sets. These findings suggest a hybrid approach—leveraging the generalizability of in-context learning alongside selective embedding-layer fine-tuning informed by historical context—may offer superior accuracy for depression-related outcomes from digital phenotyping datasets.

### 3.4 Implications for Mental Health Applications

Our findings have implications for the development of LLM-based mental health monitoring tools. The ability of LLMs to effectively integrate passive sensing data with minimal supervision or fine-tuning provides a scalable solution for monitoring changes in depressive states [42]. By identifying at-risk individuals early, such models could facilitate timely interventions, reducing the burden of mental health disorders on individuals and healthcare systems. However, given the black box nature of these algorithms and their potential to replicate existing biases in training data, these tools are best used as aides needing clinicians’ judgement and confirmation, rather than for fully automated decision-making.

There have been several prior studies using machine learning and deep learning-based methods to analyze digital phenotyping data to detect changes in depressive state. The prior study [20] uses XGBoost to examine the correlations between certain behavioral features, such as smartphone screen interaction and accelerometer data, and PHQ-9 scores. Despite these weak correlations found in the prior study, the features they find important to detect the presence and changes in severity of depressive symptoms. Similarly, our study compares multiple key behavioral features, but we go a step further by exploring nonlinear relationships and feature interactions using LLMs. The outcomes of these analyses will clarify the strengths and limitations of employing LLMs in digital phenotyping for mental health.

### 3.5 Limitations and Future Directions

While our study extends our understanding of the application of LLMs in mental health tasks, several limitations need to be addressed for a more comprehensive understanding of the models’ capabilities and their broader implications. Due to constrained computational resources and time, our study relies only on the Llama 3.1 Instruct 8B model, which has fewer than 10 billion parameters. While this model demonstrates promising results, larger LLMs with more sophisticated architectures and higher parameter counts, such as Gemma 2 27B Instruct or Llama 3.1 405B model, have demonstrated improved performance across a range of tasks and may yield even better outcomes in detecting changes in depressive state. In addition, we do not include commercial models (e.g., OpenAI’s GPT-4 or Claude 3 Opus) in our analysis due to ethical concerns around data privacy and the proprietary nature of these systems. While commercial LLMs may provide superior performance [43], the concerns around user data privacy in mental health applications are paramount.

The dataset used in this study was collected within Finland, representing a relatively homogenous cultural and geographical population. This may limit the generalizability of our findings to other regions or populations with diverse socioeconomic, cultural, or technological contexts. For instance, variations in smartphone usage patterns, access to digital infrastructure, and cultural norms around mental health reporting could influence the detection performance of LLMs in other settings. Future studies should consider cross-cultural datasets to validate the robustness of LLMs across diverse populations and to identify potential biases in model classifications.

Our dataset uses self-reported PHQ-9 scores to capture changes in depressive symptoms. All participants received a structured clinical diagnosis at intake, and PHQ-9 is widely recognized as a valid and reliable tool for tracking depressive state over time, particularly in the absence of objective biomarkers. While PHQ-9 scores are widely used in clinical and research settings, self-reported measures can be susceptible to social desirability bias, recall inaccuracies, or underreporting symptoms [44]. These limitations may affect the reliability of ground truth labels and, consequently, the model’s performance. Incorporating additional data sources, such as clinician-verified evaluations, could provide more robust labels and improve model reliability.

Our study highlights that in-context learning performance can be highly sensitive to the selection and structure of prompt examples. The decreasing detection performance observed in 3-shot and 5-shot settings highlights the importance of optimizing prompt engineering and curating high-quality, representative examples. However, designing effective prompts remains a challenging and iterative process, particularly for complex tasks such as depressive state detection. Future research should explore systematic approaches to prompt optimization, such as automated prompt generation or reinforcement learning-based methods, to maximize the utility of in-context learning [45]. This includes exploring RAG-based approaches and frameworks. Moreover, incorporating multi-modal data, such as combining smartphone sensing data with text-based self-reports or screen text [46], could enable the development of more comprehensive and accurate models.

## 4 Methods

### 4.1 Study Cohort Construction

In this study, we use a digital phenotyping dataset from a year-long longitudinal study (the Momo-Mood study) [31]. Participants are recruited in Finland from psychiatric facilities at Helsinki University Hospital, Turku University Central Hospital, and the City of Espoo Mental Health Services. The healthy controls are recruited via university email lists and voluntary healthcare staff. Diagnoses are confirmed using the Mini-International Neuropsychiatric Interview (MINI) [47] and Structured Clinical Interview-II (SCID-II) [48]. Each group has a higher proportion of female participants: the healthy control group includes 24 women and 7 men; the MDD group, 46 women and 31 men; the MDE—BD group, 18 women and 3 men; and the MDD—BPD group, 23 women and 1 man. Further details regarding participant characteristics and exclusion criteria can be found in [32].

Participants may engage in the study for up to one year, with the flexibility to withdraw anytime. The data collection process is divided into two phases: an “active phase” lasting the first two weeks, during which participants respond to 5 rounds of daily questions, and a “passive phase,” which includes biweekly PHQ-9 surveys and continuous passive sensing via smartphones. The passive sensing data comprises timestamps for communication activities (calls and texts), anonymized contact identifiers, smartphone screen events (on, off, lock, unlock), GPS location data, application usage, and battery status [20, 31]. All sensor data is collected passively from smartphone sensors. Each record includes the user’s ID, the device’s ID, and a timestamp. The study presented in this article analyzes the data collected during passive phrase using LLMs to predict PHQ-9 score changes between consecutive two-week periods.

The data are gathered using the *Niima* data collection platform [49]. The raw data is pre-processed using *Niimpy* behavioral data analysis toolbox [50]. The raw data is segmented into 6-hour bins during the day (Morning: 6:00 AM to 12:00 PM, Afternoon: 12:00 PM to 6:00 PM, Evening: 6:00 PM to 12:00 AM, Night: 12:00 AM to 06:00 AM).

These behavioral features are then aligned with PHQ-9 questionnaire responses, such that features from each two-week period are linked to the PHQ-9 score collected at the end of that period. After filtering out participants with insufficient passive data, 810 data points consisting of pairs of biweekly wearable data and PHQ-9 scores from 84 participants remain [20]. We further retain only those data points representing consecutive two-week periods, resulting in a final dataset of 666 data points.

This approach requires strict adherence to ethical standards, including safeguarding the privacy of participants and ensuring fairness in analysis to mitigate potential biases. This study was approved by the Ethical Committee of the Helsinki and Uusimaa Health District (12.09.2018; HUS/2337/2018). The project was granted a research permit by the Department of Psychiatry at the Helsinki University Hospital.

### 4.2 Task Definition and Data Labeling

This study evaluates the ability of LLMs to detect changes in depressive states using digital phenotyping data. Specifically, as shown in Figure 2, given the participant’s initial mental state, the daily behavioral sequence from the previous two weeks and the current two weeks, along with the depressive state following the previous two-week period, the model is tasked with detecting whether the depressive state will change after the current two-week period. In certain experimental settings, as shown in rows labeled with *init history* in Table 1, we additionally include historical behavioral context, i.e., the cumulative statistics of average smartphone usage to date, to assess whether including long-term behavioral patterns improves the model’s decision-making.

### Feature Selection

We select a subset of diverse features that are reported to be important for predicting changes in depressive states [20]. These features include maximum acceleration magnitude, which reflects the most significant movement or activity during a given time period; average battery level in the afternoon, indicating typical smartphone usage patterns during afternoon hours; screen-off duration during the night, representing periods of inactivity or sleep; and screen usage duration in the afternoon, highlighting engagement with the smartphone during the afternoon.

To evaluate the predictive value of these features, we define our main experimental setups by training and testing models using different combinations of the selected features. Specifically, we assess model performance when using (i) each feature individually, (ii) each pair of features, (iii) each triplet of features, and (iv) all four features together. The same model architecture and hyperparameters are applied across all experimental conditions to ensure comparability.

### Data Labeling

Figure 1 shows the process of data labeling. To capture temporal dynamics, we segment the behavioral data for each participant into two consecutive two-week periods, enabling the model to identify changes in depression state that reflect gradual changes over time. These PHQ-9 score changes are defined by comparing PHQ-9 scores at the start and end of each period, using a threshold score of 3 [51] to classify depressive states into three categories: (1) “More Depressed,” indicating a worsening condition marked by an increase in PHQ-9 score by 3 or more points from the previous week; (2) “Less Depressed,” signifying improvement, with a decrease in PHQ-9 score by 3 or more points from the previous week; and (3) “Remains stable,” reflecting no significant change, where PHQ-9 scores differed by less than 3 points between weeks. In the final dataset, there are 62 data points labeled More Depressed, 78 labeled Less Depressed, and 526 labeled Remains stable.

### 4.3 Data Splitting and Augmentation

We use the group shuffle split methodology [52] to split our dataset. This group-based splitting ensures a fair distribution of samples across the test and evaluation sets and prevents data leakage by ensuring that records from the same subject do not appear in multiple splits. The dataset is initially split into 60% training and 40% testing sets. For hyperparameter tuning, the training set is further divided into training and validation subsets in a 50:50 ratio. Once optimal hyperparameters are determined, the model is fine-tuned on the full original training set.

A significant challenge in this study is the large class imbalance in depressive state change labels, with the More Depressed and Less Depressed categories underrepresented. To address this, we apply the Synthetic Minority Oversampling Technique (SMOTE) [53] on the training set. SMOTE generates synthetic samples for the minority classes to improve their representation while preserving the underlying data distribution. Specifically, samples from the More Depressed and Less Depressed classes are resampled and augmented until each category reaches the mean number of samples across all classes.

### 4.4 Evaluation Metrics

During the evaluation phase, we assess classification performance using multiple metrics, including both macro and weighted F1 scores, alongside accuracy, which provides a straightforward measure of overall performance. Given the heavily imbalanced nature of the dataset, we primarily report the macro F1 score, as it provides a more balanced evaluation of the model’s performance across all classes, regardless of their representation.

### 4.5 LLM-based Classification Model

#### 4.5.1 Model Choice

After defining our task and building our dataset, we focus on evaluating the performance of LLMs to detect changes in the depression state. We use the Llama 3.1 8B instruct model as our base model for both in-context learning and fine-tuning settings. Llama-3 demonstrates good performance in diverse natural language tasks, including generating a summary of medical notes [54], clinical phenotype identification [55], and physical activity recognition [56].

#### 4.5.2 Zero-shot and Few-shot Prompting

One of the most exciting aspects of LLMs is their ability to solve complex problems using clear prompts tailored to specific tasks without inputting domain knowledge directly into the model. This capability enables us to evaluate LLMs by providing task-specific instructions and formatted input data.

This study starts by testing the LLMs with zero-shot prompts for detecting changes in depression state. These prompts are constructed by formatting the mobile sensor data in a structured manner, incorporating descriptions of variables and clear task instructions. We focus on the following key information:

- **Initial depressive state**: This specifies whether the participant was depressed at the time of enrollment in the study. It serves as a fixed reference point for understanding long-term changes in mental health over the study period.
- **Historical behavioral context**: This section summarizes the participant’s behavioral patterns from the time of study enrollment (init) to the beginning of the two-week reference period. This provides context for interpreting subsequent behavioral trends relative to the participant’s initial state.
- **Previous 2 weeks’ behavioral data and the mental state**: This part describes the participant’s behavioral data over the two weeks preceding the current analysis period. It includes statistical features derived from smartphone sensor data, such as the mean and standard deviation of acceleration magnitudes per day.
- **Current 2 weeks’ behavioral data**: The latest available smartphone behavioral data is included in the prompt.

One example of a structured prompt is shown in Table 2. We test two variants of prompts: one that includes historical behavioral context (corresponding to the init hist condition in Tables 1), and one without historical context (corresponding to the init condition in Tables 1).

Except for zero-shot promoting, researchers have proposed few-shot promoting techniques with LLMs, using a small number of task examples to aid in-context learning. These labeled examples are used only in the prompts sent to the model while the parameters of the LLM are frozen. We test few-shot prompting, where the model is provided with examples as demonstrations within the prompt [57]. We randomly select 1, 3, 5, 10, 15, and 25 examples (shots) as part of the prompt to explore how the number of examples influences the model’s accuracy in detecting depression state changes. These labeled examples are formatted similarly to the zero-shot prompts, but in addition, they contain structured input data and expected target outputs.

#### 4.5.3 Fine-tuning Strategies

We evaluate the performance of LLMs for detecting depressive state changes using two fine-tuning modes: parameter-efficient fine-tuning using QLoRA [34] and fine-tuning only the embedding layers [58, 59].

We apply QLoRA, where low-rank adaptation (LoRA) modules are inserted across all layers of the model [34]. Unlike some parameter-efficient approaches that only modify a few layers or components, this setup enables updates across the entire model architecture. This technique enhances the fine-tuning process’s efficiency, preserving the pre-trained model’s generalization capabilities while focusing on performance improvements. The model is loaded in 4-bit with a quantization type of nf4 (normal four-bit float) data type. The model is configured for a 32-bit paged AdamW optimizer, a batch size of 1, a warmup ratio of 0.03, gradient accumulation steps of 4, and a cosine learning rate scheduler. The QLoRA configuration includes a dropout rate of 0, a rank (r) of 64, and an alpha parameter of 16 to control the adaptation’s capacity. In embedding-only mode, the fine-tuning process updates only the embedding layer parameters, focusing on refining token representations while keeping the rest of the model frozen. By freezing the higher layers of the LLM and modifying only the embedding layers, this approach minimizes computational overhead while retaining the generalization capability of the pre-trained model. This is configured using the AdamW optimizer and similar hyperparameters as QLoRA fine-tuning mode.

### 4.6 Comparison to XGBoost Setup

We compare the in-context learning and fine-tuning classification performance with the XGboost classifier [35]. The dataset used for XGBoost contains the same features as those selected for in-context learning and fine-tuning. The data is segmented into biweekly intervals. For each time segment, a feature vector is constructed to capture sensor-based behavioral data from the preceding two weeks. This feature vector includes numerical representations of key behavioral metrics derived from passive smartphone sensor data, including average battery level in the afternoon, screen-off duration at night, screen usage duration in the afternoon, and maximum acceleration magnitude per day. To ensure consistency across models, we apply identical data augmentation techniques to each feature dataset and use the same split as in in-context learning and fine-tuning. To prevent discrepancies arising from varying feature scales, we standardize all input features before training.

Hyperparameter tuning is performed using 3-fold cross-validation [60] on the combined training and validation dataset. To identify robust and generalizable model configurations, we conduct an extensive grid search over a comprehensive set of hyper-parameters. Specifically, we vary the number of boosting rounds across 50, 100, and 200. The maximum depth of each tree is explored over a range from 3 to 7, enabling control over model complexity and overfitting. The learning rate is tuned across five values: 0.3, 0.1, 0.05, 0.01, and 0.001. To further regulate the granularity of splits, we adjust the minimum sum of instance weights needed in a child node to 1, 3, or 5. We also experiment with the subsample ratio for constructing each tree (0.1, 0.5, 0.8, and 1.0), and the fraction of features considered when growing trees (0.5, 0.8, and 1.0).

For each experimental setup—corresponding to a combination of different input features (i.e., the rows in Table 1)—the optimal hyperparameter combination is selected based on the average macro F1-score across the validation folds. Final model performance is reported on the test set using the macro F1-score to account for class imbalance. To ensure the robustness and reproducibility of results, each XGBoost model is trained and evaluated five times using different random seeds, and we report the mean performance across these runs. This rigorous tuning and evaluation protocol ensures a fair and reproducible comparison with in-context learning and fine-tuning approaches.

## Code availability

The evaluation framework used for this study can be found at: https://github.com/AsukaRoy/llm-momomood-public.

## Data Availability

The data underlying this study are not publicly accessible because of their sensitive nature, and the limitations specified in our research permit. Data access is restricted to members of the research consortium and cannot be provided to researchers outside the consortium.

## Acknowledgement

The authors gratefully acknowledge the work of their collaborators who participated in the data collection and Arsi Ikäheimonen for his valuable feedback. We also acknowledge the computational resources provided by the Aalto Science-IT project. YY also acknowledges the Foundation for Aalto University Science and Technology grant.

## Notes

### Competing Interest Statement

The authors have declared no competing interest.

### Funding Statement

This study did not receive any funding

### Author Declarations

This study was approved by the Ethical Committee of the Helsinki and Uusimaa Health District (12.09.2018; HUS/2337/2018). The project was granted a research permit by the Department of Psychiatry at the Helsinki University Hospital.

